# Authors self-disclosed use of artificial intelligence in research submissions to 49 biomedical journals: A cross-sectional study

**DOI:** 10.1101/2025.10.24.25338574

**Authors:** Isamme AlFayyad, Maurice P. Zeegers, Lex Bouter, Helen Macdonald, Sara Schroter

## Abstract

**OBJECTIVE:** To analyze the frequency of self-disclosed use of AI in research manuscripts submitted to 49 biomedical journals and to identify types of AI tools used, the tasks they assisted with, and factors associated with disclosure.

**DESIGN:** Cross-sectional study.

**SETTING:** 49 biomedical journals published by BMJ Group.

**PARTICIPANTS:** Submitting authors of 25,114 empirical research manuscripts including systematic reviews and meta-analyses, submitted between 8 April 2024 and 6 November 2024.

**MAIN OUTCOME MEASURES:** Prevalence of manuscripts with self-disclosed use of AI, types of AI tools used, the tasks they assisted with and factors associated with disclosure.

**RESULTS:** There was a total of 25,114 eligible submissions: Asia (13,505; 53.8%), Europe (6,523; 26.0%), North America (2,795; 11.1%), Africa (1,196; 4.8%), Oceania (708; 2.8%), and South America (387; 1.5%). A total of 1,431 submissions (5.7%) disclosed the use of AI. The most frequently reported AI tools used were Generative AI Chatbots (812/1431; 56.7%) and writing assistants (182; 12.7%). The majority of authors who disclosed AI use reported using it to improve the quality of their writing (1,248/1,431; 87%). Additionally, translation (107; 7.5%), generating data and output (87, 6.1%), literature searches (49; 3.4%), analyzing data (31; 2.2%), image processing or analysis (49; 3.4%), code writing (15; 1.0%), and managing references (14; 0.9%) were mentioned as tasks AI assisted with.

Authors from South America (OR=1.75; 95%-CI: 1.22-2.49) and Europe (OR=1.28; 95%-CI: 1.14-1.45) were significantly more likely to disclose AI use than those from Asia. Conversely, each additional author reduced disclosure odds by 1% (OR=0.99, 95%-CI: 0.97-0.99). Acceptance rate, impact factor, type of journal, and peer review model) were not associated with AI use disclosure.

**CONCLUSIONS:** We found that only a small proportion (5.7%) of submitting authors disclosed AI use, which is substantially lower than proportions reported in surveys. Improving the quality of writing was the primarily task AI assisted with and AI Chatbots were the most commonly disclosed tool. Authors may be uncertain about what AI use requires disclosure or may be hesitant to declare it.

**What is already known on this topic:**

▪ Artificial intelligence is increasingly used in the conduct of scientific research and writing of publications and the way it is used is changing rapidly.
▪ Leading publishers and editorial organizations dedicated to promoting integrity and best practices in academic publishing have issued policies requiring authors to disclose all AI use in their content.
▪ Previous surveys show that a substantial proportion of researchers (28% to 76%) say they have used AI in their research.

**What this study adds:**

▪ This study of a process for mandatory declaration of AI use in research manuscripts submitted to 49 biomedical journals found a prevalence of only 5.7%, which is substantially lower than use reported in surveys.
▪ Improving the quality of writing was the most common reported task performed by AI, and _g_enerative AI Chatbots are the most frequently disclosed AI tool used.

**How this study might affect research, practice or policy:**

▪ Current mandatory questions on AI use may have only limited value and other measures are needed to improve declaration and to support detection of undisclosed use.

## INTRODUCTION

Artificial intelligence (AI), including Generative AI (GAI), has rapidly emerged as a powerful tool in scientific research and publication [**1**]. AI encompasses a broad spectrum of tools, ranging from machine learning algorithms for data analysis, GAI tools, such as natural language processing (NLP) systems for text mining and summarization, to computer vision for image recognition and analysis [**2**]. GAI tools, such as Chatbots (e.g., ChatGPT, Gemini, Claude) are capable of generating scientific content that closely resembles or even rivals human capacity in terms of accuracy, answering complex questions, and speed when performing specific tasks [**3**]. GAI tools trained on large datasets enable researchers to perform research-related tasks and influence the way scholarly information is accessed, organised, analysed, and disseminated, thus reforming scientific research and the publication of its findings [**4**].

Leading academic publishers and editorial organisations have issued recommendations or position statements to support editors in developing AI policies [**5-9**]. These recommendations are aimed at the transparency of reporting AI use, specifically disclosure of the AI tool name, version, and what task the AI was used for. All these organizations make it clear that authors remain responsible for content generated by AI [**5-9**] and that therefore AI should not be listed as an author, because it “cannot be responsible for the accuracy, integrity, and originality of the work” [**9**]. The European Association of Science Editors (EASE) recommends that any details regarding the application of AI within a manuscript be articulated in relevant manuscript sections [**7**]. This encompasses, but is not restricted to, data collection, data cleaning, code generation, analysis, interpretation, visualisation, and the processes of writing and language editing [**7**]. BMJ Group journals will consider for publication manuscripts where content has been influenced by AI’s technologies, provided that the authors explicitly and thoroughly describe the application of these technologies in the contributorship statement to allow proper evaluation of AI role in research by editors, reviewers, and readers [**10**]. The policy allows flexibility for the editors to judge that the use was reasonable and to publish the work where appropriate. However, in other cases, editors may judge that the AI use was not suitable or sufficiently reliable, and they may choose to reject the content from the journal.

Despite efforts to promote transparency regarding AI use, the extent of authors’ compliance with such disclosure requirements remains uncertain. Current estimates of AI use rely entirely on self-reported surveys with limited samples [**11-16**]. These surveys have indicated widespread use of AI tools among researchers (28% up to 76%). The findings suggest that AI use has become common practice in research. However, a recent review of manuscripts submitted to JAMA Network journals showed that only 2.7% disclosed AI use [17]. In general, there is little guidance for authors on what types of use need to be disclosed. Resnik and Hosseini recommend mandatory disclosure when AI tools are intentionally and substantially used in research. They defined intentional AI use in research as when “one purposefully employs a unique AI tool to conduct a specific task”, while substantial contribution concerns instances where AI makes decisions that impact research results or when AI-generates or synthesizes content directly influencing research outcomes and publication [**18**].

In 2023, BMJ Group developed an AI use policy, and on April 8, 2024, implemented a mandatory process to support self-declaration using questions on AI use in ScholarOne (its manuscript submission and tracking portal) for completion by submitting authors (Box 1). This enables editors, reviewers, and readers to evaluate whether the AI use is suitable and reasonable [**10**]. In this context, our study aims to provide a large-scale analysis of self-disclosed AI use in original research manuscripts submitted to 49 biomedical journals published by BMJ Group, examining the frequency of AI use, types of AI tools, tasks these assist with, and factors associated with disclosure.

**Box.1:**
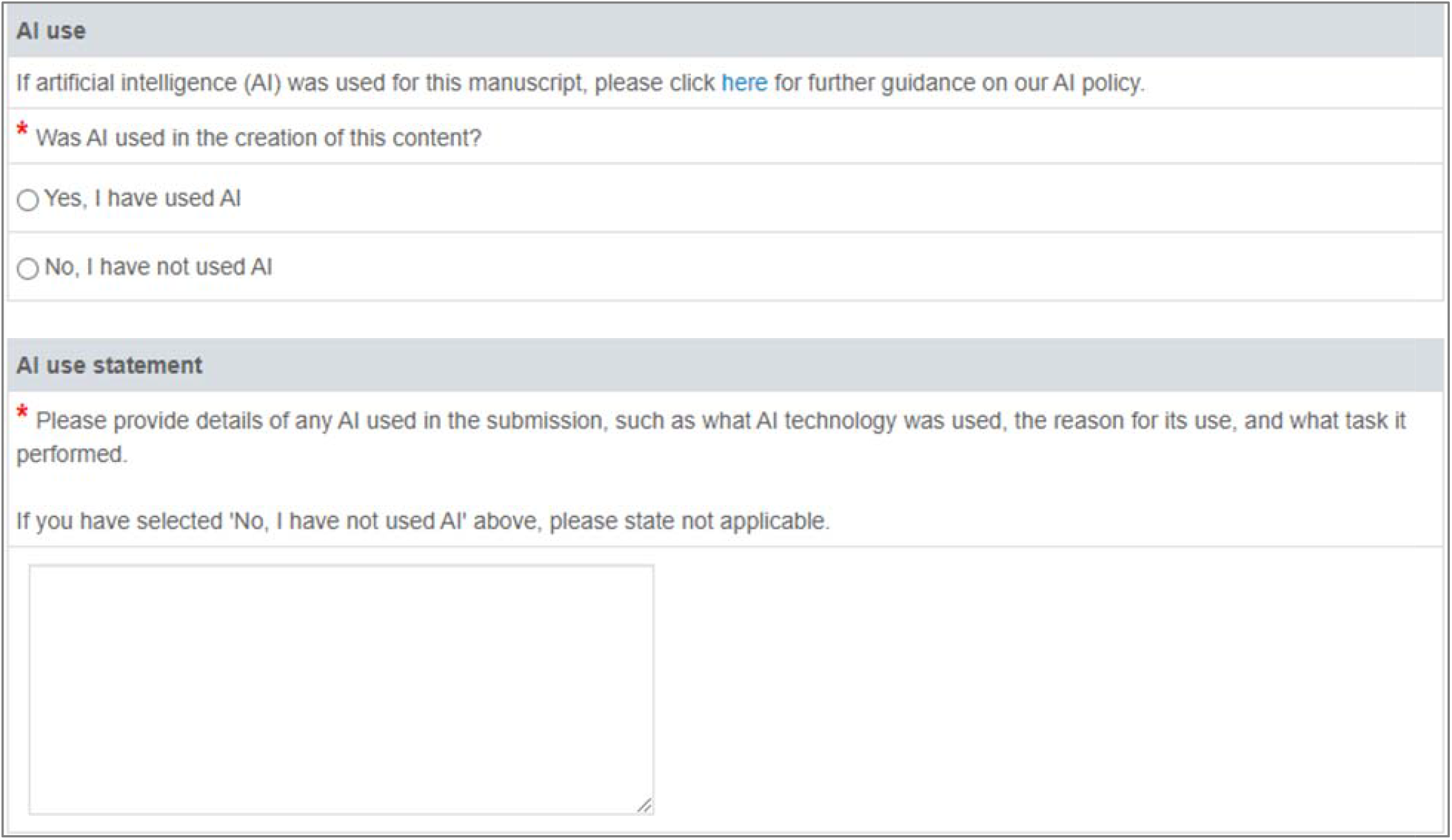
Mandatory questions introduced in BMJ Group journals’ manuscript submission systems for disclosure of AI use.

## METHODS

### Study design

A cross-sectional analysis of authors’ responses to mandatory AI use disclosure questions in research manuscripts submitted to 49 biomedical journals published by BMJ Group.

### Participants and data source

Using the journals’ ScholarOne manuscript tracking systems, we identified and included the first version of manuscripts reporting on empirical research, including systematic reviews and meta-analyses, submitted to 49 biomedical BMJ Group journals between 8^th^ of April and 6^th^ of November 2024. Manuscripts of other types, such as commentaries, case reports/series, opinion pieces, editorials, and letters were excluded from analysis.

### Data Collection and outcomes

Characteristics of manuscripts, authors and journals were extracted from ScholarOne manuscript tracking systems and journals’ websites. We extracted the submitting authors’ geographical region of affiliation (Europe, North America, South America, Asia, Africa, Oceania) and the total number of authors of the manuscripts. We also extracted manuscript ID, date of initial submission, manuscript title, abstract, and research subject area. Data at the journal level included journal type (general medicine vs specialised), type of peer review model (open peer review, single blinded, double blinded), impact factor (categorised as No impact factor, ≤5, >5-10, >10-20, >20), and the current journals’ acceptance rate reported on the journals’ websites. Primary outcomes were: (1) the frequency of self-disclosed AI use, (2) types of AI tools used, and (3) the tasks they assisted with.

In the absence of consensus or a published international standard for categorizing AI tools in research as of early 2025, submitting authors’ free-text disclosures were systematically reviewed and classified based on their function□based taxonomy that grouped tools according to their primary purpose within the manuscript. If the submitting author’s description was not clear or did not specify the tool’s main function, then we looked up official documentation (such as the tool’s website) to determine what the tool was designed to do and how it is typically used. Categories included: AI Chatbots, Writing Assistants, Visual/Image Processing Tools, Evidence Synthesis Tools, Predictive Analytics Models, AI□powered Translators, AI□powered Data Analysis Tools, and Other Specialized Tools (including niche applications used by a very small proportion of respondents). Cases where no tool type was specified were classified as type not disclosed.

### Data analysis

Descriptive statistics (frequencies, proportions, means, standard deviations) were used to summarise characteristics of manuscripts, authors and journals, as well as the frequency of self-disclosed AI use, types of AI tools used, and the tasks AI was used for. Chi-square tests compared the proportions of AI users versus non-users across characteristics of manuscripts, authors, and journals. Independent sample t-test compared differences in means (number of authors, journal acceptance rate) between manuscripts declaring AI use versus not. Binomial logistic regression was performed to identify factors associated with self-disclosed AI use after controlling for other factors. Six factors were included in the model: journal acceptance rate, impact factor, peer review model, journal specialty, submitting author’s region, and number of authors. Results presented as odds ratios (OR) with 95% confidence intervals (CI). Level of statistical significance was set at <0.05.

### Patient and Public Involvement

We did not involve patients or the public in the design, conduct, or reporting of this research, as it did not involve clinical data or patient participants.

### Ethical considerations

All identifying author data were removed from the sample and the data were stored in a shared BMJ Google Drive. Ethical approval for the study was obtained from the research ethics committee of Maastricht University (Approval Number: FHML-REC/2025/014). The research protocol was registered in open science framework (OSF) before we started the analysis and is available at osf.io/uvqms. Prior to initiating the study, a research agreement was signed between the primary author (IA) and BMJ Group and the research was conducted as part of BMJ’s quality improvement program. All submitting authors are informed on submission that BMJ has an ongoing programme of research and are offered the possibility to opt out.

## RESULTS

### Sample

A total of 25,114 eligible submissions were analysed (Table 1). The mean journals’ acceptance rate was 25.6%±11.2%, and the mean number of authors per submission was 8.9±6.5. Most manuscripts 15,289 (60.9%) were submitted to single anonymized peer review journals and 14,858 (59.2%) of submissions were to specialized medical journals. Submissions were received from 140 countries. Asia accounted for over half of submissions 13,505 (53.8%), followed by Europe 6,523 (26.0%), North America 2,795 (11.1%), Africa 1,196 (4.8%), Oceania 708 (2.8%), and South America 387 (1.5%). More than half were submitted to journals with an impact factor less than 5.

**Table 1.**
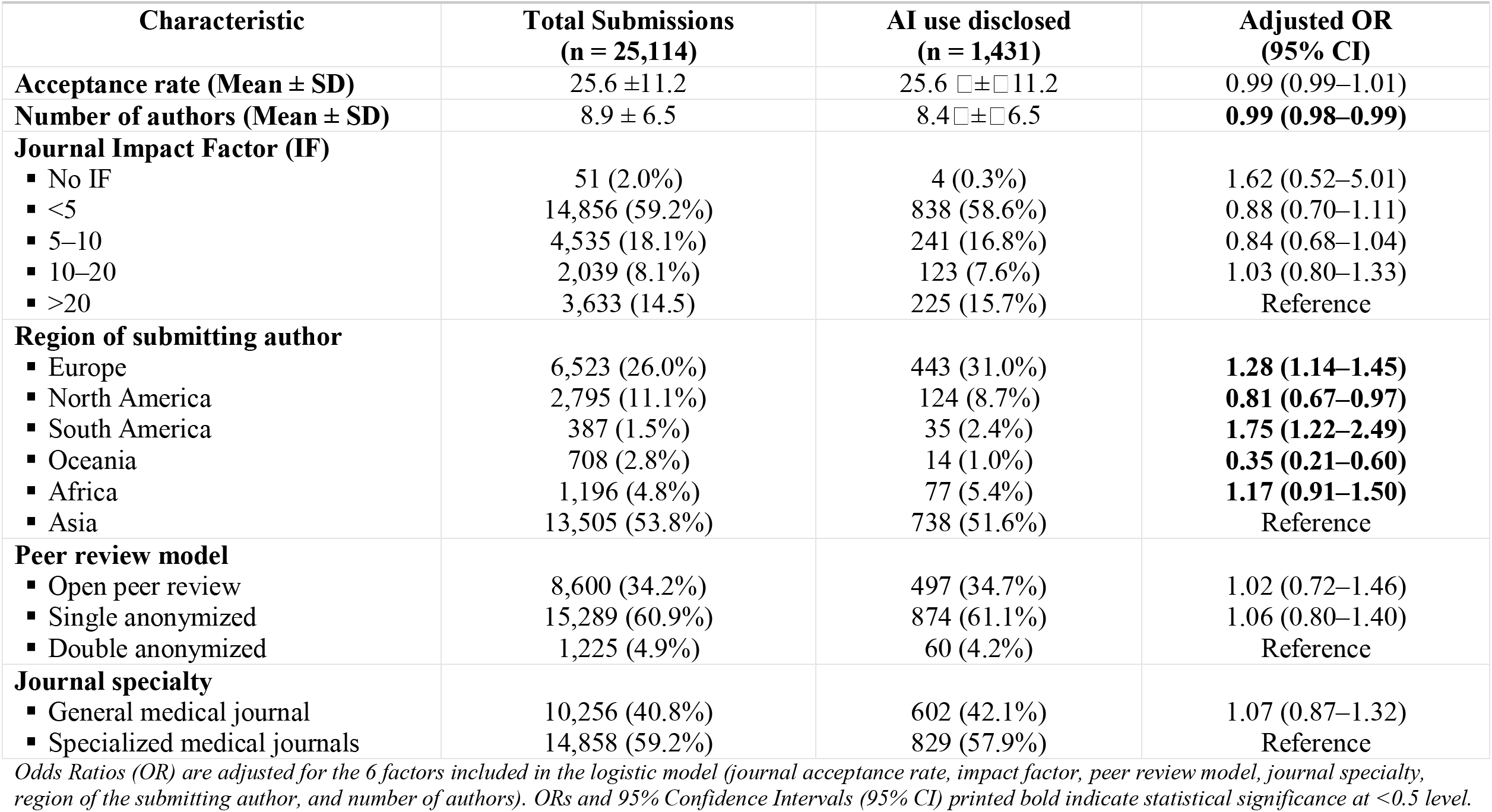
Descriptive analysis of submissions disclosing AI use vs. not and factors associated with AI disclosure.

**Table 2.** Reporting prevalence of types AI tool used.

| **Table 2.** Reporting prevalence of types AI tool used |  |
| --- | --- |
| **Type** | **n (%)** |
| AI Chatbots | 812 (56.7%) |
| Writing assistant tools | 182 (12.7%) |
| Visual/image Processing Tools | 38 (2.7%) |
| Evidence Synthesis Tools | 28 (2%) |
| Predictive Analytics Models | 21 (1.5%) |
| AI powered translators | 19 (1.3%) |
| AI-powered data analysis tools | 9 (0.6%) |
| AI-powered markdown editors | 7 (0.5%) |
| Automatic Speech Recognition Systems | 4 (0.3%) |
| Other | 7 (0.5%) |
| Type not disclosed | 449 (31.4%) |

*The sum of percentages exceeds 100%, because multiple AI tools may have been reported per manuscript*

### Prevalence of self-disclosed use of AI and factors associated with AI use disclosure

Of the 25,114 eligible submissions, 1,431 (5.7%) disclosed the use of AI. The number of authors on the authorship byline was slightly lower in manuscripts for which AI-use was disclosed (8.4□±□6.5 versus 8.9□±□6.5). Manuscripts with disclosed AI use were proportionally more likely to have a submitting author from Europe (31.0% vs. 25.7%) and South America (2.4% vs. 1.5%), while manuscripts with AI use not disclosed were more likely to have a submitting author from Asia (53.9% vs. 51.6%) and Oceania (2.9% vs. 1.0%). The binominal logistic regression analysis identified two factors associated with the likelihood of disclosing AI use. A small inverse association was observed with the number of authors (OR 0.99, 95% CI: 0.98–0.99), indicating that manuscripts with more authors were slightly less likely to report AI use. Submitting authors from South America (OR 1.75, 95% CI: 1.22–2.49) and Europe (OR 1.28, 95% CI: 1.14–1.45) had significantly higher odds of disclosing AI use compared to those from Asia.

### Types of AI tools used

Among the 1,431 submissions that disclosed AI use, the tools varied considerably. AI Chatbots were reported in 812 submissions (56.7%). Writing assistant tools were reported in 182 submissions (12.7%). Other AI types, including visual/image processing tools, evidence synthesis tools, predictive analytics models, AI-powered translators, and automatic speech recognition systems were reported for fewer than 3% of submissions each. For an additional 7 submissions (0.5%), the use of other AI tools not fitting our function-based categorization was reported. Notably, for 449 (31.4%) submissions, the AI tool used was not disclosed.

### Tasks AI tools assisted with

Table 3 reveals how self-disclosed AI use is being utilized to assist with various tasks. The predominant use of AI was to improve the quality of writing (87%, 1248). Other tasks supported by AI were considerably less common, including translation (7.5%), generating data and output (6.1%), literature searches (3.4%), and analysing data (2.2%). Additionally, 0.9% of respondents did not disclose the specific task for which AI was used, and 1.3% indicated “other” tasks.

**Table 3.** Prevalence of tasks AI is used for.

| Tasks | n (%) |
| --- | --- |
| To improve the quality of writing | 1248 (87%) |
| To translate text | 107 (7.5%) |
| To generate data and output | 87 (6.1%) |
| To search the literature | 49 (3.4%) |
| To analyse data | 31 (2.2%) |
| To process or analyse images | 49 (3.4%) |
| To write code | 15 (1%) |
| To manage references | 14 (0.9%) |
| To detect originality of text | 12 (0.8%) |
| Other | 18 (1.3%) |
| Task not disclosed | 13 (0.9%) |

*The sum of percentages exceeds 100%, because authors may have reported more than 1 task.*

## DISCUSSION

Our large-scale analysis of mandatory self-disclosed AI use in research manuscripts submitted to BMJ Group biomedical journals found that among 25,114 submissions, only 5.7% reported AI use. The most frequently reported AI tools used were Generative AI Chatbots and writing assistants. The majority of authors who disclosed AI use reported using it to improve the quality of their writing. It was also used for translation, generating data and output, literature searches, analyzing or collecting data, image processing, code writing, and managing references. Authors from South America and Europe were significantly more likely to disclose AI use than those from Asia. Manuscripts with more authors were less likely to disclose AI use.

The prevalence of self-disclosed AI use in our study is substantially lower than prevalence estimates from surveys [**11-14**]. However, BMJ Group’s mandatory disclosure captures AI use prevalence at the individual manuscript level, whereas surveys assess broader researcher-level use, so we would expect some differences in prevalence. Reported prevalence rates in surveys vary widely: 76% of 2,345 researchers in the Oxford University Press survey [**11**], over 50% of 3,000 researchers in the Elsevier survey [**12**], 28% of 5,000 researchers in the Nature survey with 18% nondisclosure [**13**], 45% of 4,946 researchers in Wiley’s ExplanAItions survey [**16**], 44.5% of 2,125 researchers in Ng et al.’s study [**14**], and 58.5% of Chinese scholars in a recent survey, although only 29% had disclosed such use in submitted manuscripts [**19**].

Consistent with our findings that 5.7% of submissions to BMJ Group journals disclosed AI use; a recent study of 82,829 manuscripts submitted to JAMA Network journals reported 2.7% contained declarations of AI use, increasing from 1.6% to 4.2% across 2023–2025 [**17**]. Another recent submissions analysis, this time in the cancer research field, used automated AI detection software and flagged AI text in up to 23% of abstracts in 2024 [**20**]. Collectively, these results highlight the disparity between self-disclosed and actual use of AI.

Several factors may contribute to underreporting of AI use. Our findings cannot determine whether this underreporting actually exists and whether it stems from deliberate omission, or non-adherence with editorial policies, or uncertainty over what AI use needs to be disclosed. The concept of uncertainty has been examined by Resnik and Hosseini (2025), who classified AI disclosure as mandatory, optional, or unnecessary. This classification underscores the ongoing ambiguity about what constitutes intentional and substantial AI use—an ambiguity that may ultimately influence authors’ decisions regarding disclosure [**18**].

GAI Chatbots were the most frequently disclosed tools in our study (56.7%), exceeding rates reported in recent surveys, including Oxford University Press (43%), Elsevier (25%), Wiley (35%), and Ng et al. (44.5%) [**11–12**], while Nature (2024) similarly identified ChatGPT as the most widely used [**13**]. This higher prevalence may reflect recent adoption of AI, stricter BMJ disclosure policies, differences in authors reporting behavior, or that these surveys captured researchers’ general use.

AI writing assistant tools (e.g., Grammarly, Quillbot, Hemingway Editor) for supporting grammar correction, language refinement, clarity enhancement, and structural consistency, helping authors produce more polished manuscripts are increasingly common among researchers [**11–16,18,21,22**]. In our study, AI writing assistant tools were the second most disclosed tools (12.7%), comparable to Ng et al. (12.6%) but lower than the 19–38% reported elsewhere [**11,13,16**].

By contrast, disclosures of AI use for complex research activities-data analysis, code generation, or image processing - were rare in our study. This may reflect cautious application of AI to core scientific tasks or underreporting due to uncertainty over what constitutes acceptable use, limited adoption or hesitancy to disclose. Overall, the predominance of Chatbots and writing assistants highlights that AI in research is currently applied more to enhancing writing quality than to conducting primary scientific work.

Some authors reported AI use without specifying the tool or task limiting our insight to estimate prevalence of each tool or task with rigor. Overall, the predominance of AI-assisted writing underscores its current role as a facilitator of manuscript preparation.

Unlike previous surveys [**11–16**], our study provides continent-level estimates of AI disclosure and reveals notable regional variation. Authors from South America and Europe were significantly more likely to report AI use than those from Asia, even after adjusting for other factors, which may reflect differences in reporting culture, AI access and training, or local institutional policies. Additionally, disclosure was inversely associated with the number of authors, possibly due to coordination challenges in larger teams.

The AI tool classification in this study differs substantially from consensus-driven frameworks recently proposed by Schoenenberger et al and the STM Recommendations for a classification of AI Use [**23,24**]. Our classification was empirically grounded, based on a function-oriented taxonomy developed from reviewing real-world author disclosures in BMJ Group journal submissions. Unlike the activity-focused categories proposed for harmonizing editorial policy, our classification emphasized the actual purpose and context in which each tool was used (e.g., Chatbots, writing assistants, translators). When disclosures were ambiguous, official tool documentation was checked, while unreported AI tools were marked as “type not disclosed.” This distinction makes our classification more reflective of current author behaviors and the nuanced challenges in self-reporting AI use, rather than being theoretical classifications intended primarily for future policy guidance.

### Strengths and Limitations

The main strength of this study is its large sample size (all eligible submitted manuscripts) and broad representation across 49 BMJ Group biomedical journals, encompassing over 25,000 first submissions of empirical research from diverse global regions. By using real-world data from mandatory AI disclosures at submission, the study provides a more current measure of AI use in scholarly publishing than previous survey-based estimates. However, several limitations should be noted. First, the study relies on self-disclosure, which may be affected by selective underreporting, misunderstanding, or omission, potentially creating gaps in the data. Second, the analysis is limited to BMJ Group biomedical journals, which may reduce generalizability to other publishers with different editorial policies or different journals’ content scope. Finally, the manuscript tracking systems do not capture author characteristics such as age, gender, career stage, discipline, or academic role, limiting exploration of their potential influence.

### Implications for Future Research, Practice, and Policy

The findings of this study provide a foundation for further research to systematically evaluate discrepancies between self-reported and software-detected AI use in BMJ Group manuscript submissions. Future studies should explore cognitive and cultural factors influencing authors’ disclosure behaviors, and longitudinal research is needed to track trends in AI use disclosure over time. Additionally, the potential impact of disclosed AI use on publication outcomes warrants investigation. Based on our findings, we recommend that journals consider implementing a more structured AI disclosure question during manuscript submission, including dropdown menus specifying AI tools and tasks. This may improve the quality or extent of disclosure by prompting authors to consider particular steps of their research. This could be complemented with validated automated AI-detection tools, if available, particularly those integrated into editorial workflows. Clearer policies defining permissible AI use may help encourage disclosure of clearly permissible activities by authors, in addition to and emphasizing potential consequences of non-disclosure such as rejection and or possible retraction of work.

Such a complementary approach would enhance consistency of AI use reporting, minimize the risk of undisclosed AI use, and safeguard research integrity.

## CONCLUSIONS

Despite increasing use of AI tools in research and mandatory disclosure policies, self-disclosure of AI use is low, possibly suggesting substantial (selective) underreporting that undermines transparency and accountability in scientific publishing. Future research should explore barriers to AI disclosure, evaluate the effectiveness of policy interventions, and assess the impact of AI use on research quality and integrity.

## Data Availability

The anonymized dataset is publicly available at osf.io/uvqms.

https://osf.io/

## Contributors

IA, MPZ, LB, HM, and SS were involved in the study concept and design. SS was responsible for data collection and extraction. IA did the data analysis and IA, MPZ, LB, HM, and SS interpreted the data. IA wrote the initial draft manuscript and managed the versions. All authors were involved in critical revision of multiple drafts and approved the final version. The corresponding author attests that all listed authors meet authorship criteria and that no others meeting the criteria have been omitted intentionally. All contributors have been invited to be included in the list of authors, and their choice has been fully respected.

## Patient consent for publication

Not applicable.

## Ethics approval

Ethical approval was obtained from the research ethics committee of Maastricht University (Approval Number: FHML-REC/2025/014).

## Data availability statement

The anonymized dataset is publicly available at osf.io/uvqms.

## Declaration of interest

SS and HM are full time employees of BMJ Group. This research is part of an ongoing PhD collaboration between BMJ Group and the team Meta-Research at Maastricht University (UM) on the responsible conduct of publishing scientific research. BMJ

Group is a wholly owned subsidiary of the British Medical Association. UM is a public legal entity in the Netherlands. This study is part of Isamme AlFayyad self-funded BMJ Group/UM PhD trajectory. No exchange of funds has taken place for this research project. All authors express their own opinions and not necessarily that of their employers.

## Notes

### Funding Statement

This study did not receive any funding

